# Single View Echocardiographic Analysis for LVOT Obstruction Prediction in Hypertrophic Cardiomyopathy: A Deep Learning Approach

**DOI:** 10.1101/2025.03.10.25323713

**Authors:** Jiesuck Park, Jiyeon Kim, Jaeik Jeon, Yeonyee E. Yoon, Yeonggul Jang, Hyunseok Jeong, Seung-Ah Lee, Hong-Mi Choi, In-Chang Hwang, Goo-Yeong Cho, Hyuk-Jae Chang

## Abstract

**Background:** Accurate left ventricular outflow tract obstruction (LVOTO) assessment is crucial for hypertrophic cardiomyopathy (HCM) management and prognosis. Traditional methods, requiring multiple views, Doppler, and provocation, is often infeasible, especially where resources are limited. This study aimed to develop and validate a deep learning (DL) model capable of predicting severe LVOTO in HCM patients using only the parasternal long-axis (PLAX) view from transthoracic echocardiography (TTE).

**Methods:** A DL model was trained on PLAX videos extracted from TTE examinations (developmental dataset, n=1,007) to capture both morphological and dynamic motion features, generating a DL index for LVOTO (DLi-LVOTO, range 0-100). Performance was evaluated in an internal test dataset (ITDS, n=87) and externally validated in the distinct hospital dataset (DHDS, n=1,334) and the LVOTO reduction treatment dataset (n=156).

**Results:** The model achieved high accuracy in detecting severe LVOTO (pressure gradient≥ 50mmHg), with area under the receiver operating characteristics curve (AUROC) of 0.97 (95% confidence interval: 0.92-1.00) in ITDS and 0.93 (0.92-0.95) in DHDS. At a DLi-LVOTO threshold of 70, the model demonstrated a specificity of 97.3% and negative predictive value (NPV) of 96.1% in ITDS. In DHDS, a cutoff of 60 yielded a specificity of 94.6% and NPV of 95.5%. DLi-LVOTO also decreased significantly after surgical myectomy or Mavacamten treatment, correlating with reductions in peak pressure gradient (p<0.001 for all).

**Conclusions:** Our DL-based approach predicts severe LVOTO using only the PLAX view from TTE, serving as a complementary tool, particularly in resource-limited settings or when Doppler is unavailable, and for monitoring treatment response.

## 1. INTRODUCTION

Hypertrophic cardiomyopathy (HCM) is one of the most common genetic cardiomyopathies, with an estimated prevalence of 1 in 200 to 1 in 500.^1^ The phenotype of HCM is highly heterogeneous, ranging from asymptomatic presentation to severe outcomes such as sudden cardiac death (SCD). Among the various factors influencing symptoms and prognosis, left ventricular outflow tract (LVOT) obstruction (LVOTO) plays a pivotal role.^1,2^ When LVOTO becomes severe, it not only contributes to symptoms such as exertional dyspnea, chest pain, and syncope but also serves as the key determinant in guiding clinical management, including pharmacological treatment, septal reduction intervention, or surgery.^2^ Therefore, accurate identification of LVOTO is critical in the clinical evaluation of HCM patients.

Transthoracic echocardiography (TTE) is the primary imaging modality for diagnosing and evaluating HCM.^2^ LVOTO is typically assessed through TTE using Doppler imaging and multiple echocardiographic views, often incorporating provocation maneuvers.^2^ However, these assessments require substantial expertise in image acquisition and interpretation, are time-intensive, and are subject to technical variability, interobserver variability, and patient cooperation. These limitations can hinder their feasibility, especially in acute care settings using handheld devices, in resource-limited community hospitals, or in patients unable to tolerate provocation. Among TTE views, the parasternal long axis (PLAX) view is one of the most fundamental, offering critical insights into cardiac anatomy and dynamic function. Therefore, leveraging this single, widely available view for automated prediction of LVOTO could improve accessibility and maintain clinical accuracy, even in settings where expert evaluation is limited or advanced Doppler-equipped systems are unavailable.

Recent advancements in deep learning (DL) have demonstrated significant potential in automating and enhancing medical image interpretation, and TTE is no exception. DL algorithms can identify complex patterns in imaging data that may elude human observers, thereby improving diagnostic accuracy and efficiency. For instance, efforts have been made to accurately assess conditions such as aortic stenosis (AS) and diastolic dysfunction using limited 2-dimensional (2D) TTE images, even without Doppler input.^3–7^ Despite these advances, the application of DL for predicting LVOTO using single-view TTE in HCM patients remains underexplored. A recent attempt utilized the apical four-chamber (A4C) view to predict LVOTO through a DL-based model.^8^ However, this single-view approach is fundamentally limited by the A4C’s indirect visualization of the LVOT, resulting in suboptimal performance that may restrict its clinical applicability.

In the present study, we aimed to develop a hybrid spatiotemporal network that learns directly from resting 2D PLAX TTE videos, which provide a clear visualization of the LVOT, to predict the presence of severe LVOTO in HCM patients without the need for Doppler input or provocation maneuvers. To achieve this, our framework integrates global B-mode spatial context with multi-resolution M-mode features, enabling the construction of a comprehensive spatiotemporal representation of LVOTO dynamics. Specifically, we designed a novel, fully automated multi-slice M-mode generation framework derived directly from PLAX 2D video data, eliminating the need for manual placement or predefined anatomical landmarks. This multi-slice M-mode approach dynamically tracks key anatomical structures throughout the cardiac cycle, capturing subtle hemodynamic cues that are often only observed during provocation maneuvers. Our model’s ability to synchronize spatial and temporal information allows for the detection of flow abnormalities and structural changes in real time, enhancing both interpretability and diagnostic precision. This study presents the development and validation of our DL-based model and evaluates its predictive performance for severe LVOTO. Additionally, we assess its potential as a practical complementary tool to conventional Doppler-based assessments, which often require multiple TTE views with provocation maneuvers, offering a streamlined method for LVOTO detection, particularly valuable in resource-limited settings and for less experienced operators.

## 2. METHODS

### 2.1. Study Population and Data Sources

The DL-based model presented in this study was developed and validated using the Open AI Dataset Project (AI-Hub) dataset, an initiative supported by the South Korean government’s Ministry of Science and Information and Communication Technology (ICT).^5,6,9–13^ This dataset comprised 30,000 echocardiographic examinations retrospectively collected from five tertiary hospitals between 2012 and 2021, encompassing a wide range of cardiovascular diseases, including HCM. From this dataset, we identified cases categorized as HCM and applied the following inclusion criteria: (1) availability of echocardiographic reports, (2) a clinical diagnosis of HCM established by identifying a maximal end-diastolic wall thickness ≥15mm in any segment of the left ventricle (LV), with other potential causes of hypertrophy excluded, and (3) TTE in which the presence or absence of LVOTO had been assessed by Doppler-based pressure gradient (PG) measurements. While Doppler data were necessary to establish ground truth (GT) for LVOTO classification, they were not used as model input. Furthermore, no examinations were excluded based on image quality, as we aimed to capture typical variations observed in real-world clinical TTE images.

To develop the DL-based LVOTO prediction algorithm, we created the Developmental Dataset (DDS), consisting of 722 patients with 1,007 TTE examinations, which were divided into training, validation, and test datasets in an 8:1:1 ratio, comprising 578, 71, and 73 patients (809, 111, and 87 TTE examinations, respectively). (**Figure 1**) TTE data from Severance Hospital were excluded from the DDS and designated as the Distinct Hospital Dataset (DHDS) for independent external tests. The DHDS included 573 patients with 1,334 TTE examinations, serving as external dataset to evaluate model generalizability. Both the DDS and DHDS explicitly excluded any TTE data from patients who had undergone LVOT gradient reduction treatments, including surgical myectomy or Mavacamten therapy, to avoid potential confounding effects during model development and validation. Additionally, we collected pre- and post-treatment TTE data from obstructive HCM patients who underwent LVOT gradient reduction treatment at Seoul National University Bundang Hospital and Severance Hospital. This dataset comprises 17 patients with 112 TTE examinations from those who underwent surgical myectomy and 13 patients with 44 TTE examinations from those treated with Mavacamten. To eliminate any risk of data leakage, these patients were entirely excluded from the DDS, regardless of when their TTE examinations were performed.

**Figure 1.**
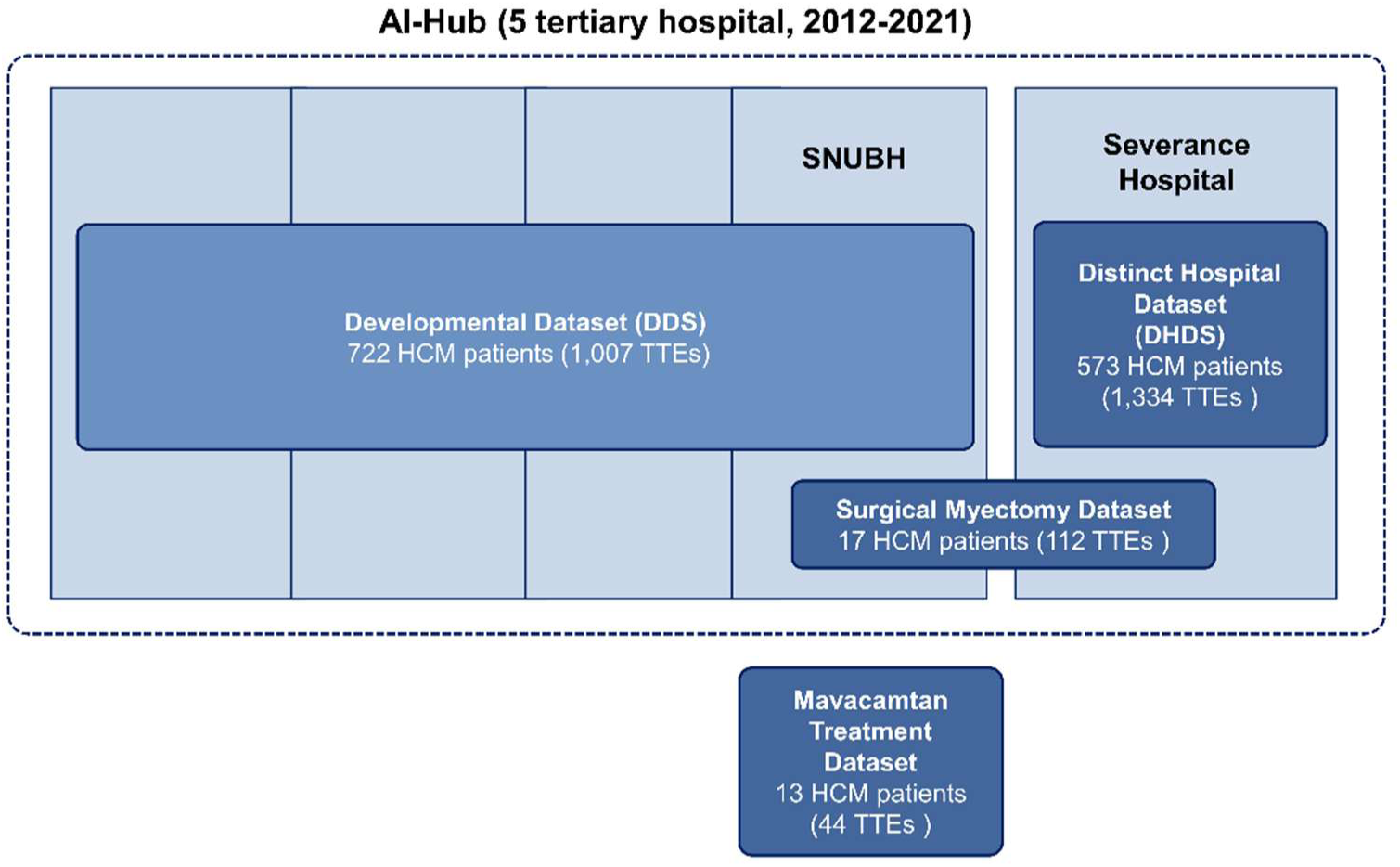
Overview of Dataset Composition for DL-Based LVOTO Prediction Model Development and Validation. This figure illustrates the composition of datasets used for the development and validation of the DL-based LVOTO prediction model. The Developmental Dataset (DDS) was derived from multiple tertiary hospitals, excluding data from Severance Hospital, which was designated as the Distinct Hospital Dataset (DHDS) for independent external validation. Additionally, we collected baseline and follow-up TTE data from obstructive HCM patients who underwent surgical myectomy or Mavacamten treatment, forming the Surgical Myectomy Dataset and Mavacamten Treatment Dataset, respectively. Each dataset is independent, and no patients were shared across datasets to prevent data leakage and ensure unbiased model training and evaluation. DL, deep learning; LVOTO, left ventricular outflow tract obstruction; SNUBH, Seoul National University Bundang Hospital.

The study protocol was approved by the Institutional Review Boards of all participating institutions, with a waiver of informed consent granted due to the retrospective study design. All clinical data were fully anonymized prior to analysis. The study was conducted in accordance with the principles outlined in the Declaration of Helsinki (2013).

### 2.2. TTE Acquisition and Interpretation

All TTE studies were conducted by trained echocardiographers or cardiologists and interpreted by board-certified cardiologists specializing in echocardiography as part of routine clinical care. All TTE examinations included in this study were standard TTEs; exercise or pharmacological stress tests were not included. However, provocation maneuvers that can be performed during standard TTE, such as the Valsalva maneuver, were included to evaluate LVOTO. For the assessment of LVOTO, continuous-wave Doppler was used in the apical 3-chamber or 5-chamber view to measure the LVOT peak velocity (V_max_, m/sec). The Bernoulli equation (PG = 4 × Vmax²) was then applied to calculate the LVOT peak PG. If a Valsalva maneuver successfully induced or exacerbated LVOTO, the peak PG measured during Valsalva was used for classification. Conversely, if the Valsalva maneuver did not lead to any measurable increase in LVOTO, the resting PG was used instead. LVOTO was classified based on the LVOT peak pressure gradient (PG): a peak PG ≥ 30 mmHg was defined as significant LVOTO, and a peak PG ≥ 50 mmHg was defined as severe LVOTO.^2,14^

### 2.3. Model Development

We developed a novel DL model to predict severe LVOTO using single-view, 2D PLAX TTE videos acquired at rest, eliminating the need for Doppler assessments. The model was trained to infer the presence of severe LVOTO as determined by Doppler assessment. It was explicitly designed to capture both morphological (i.e., spatial) and motion (M-mode) features critical for LVOTO assessment. (**Figure 2**)

**Figure 2.**
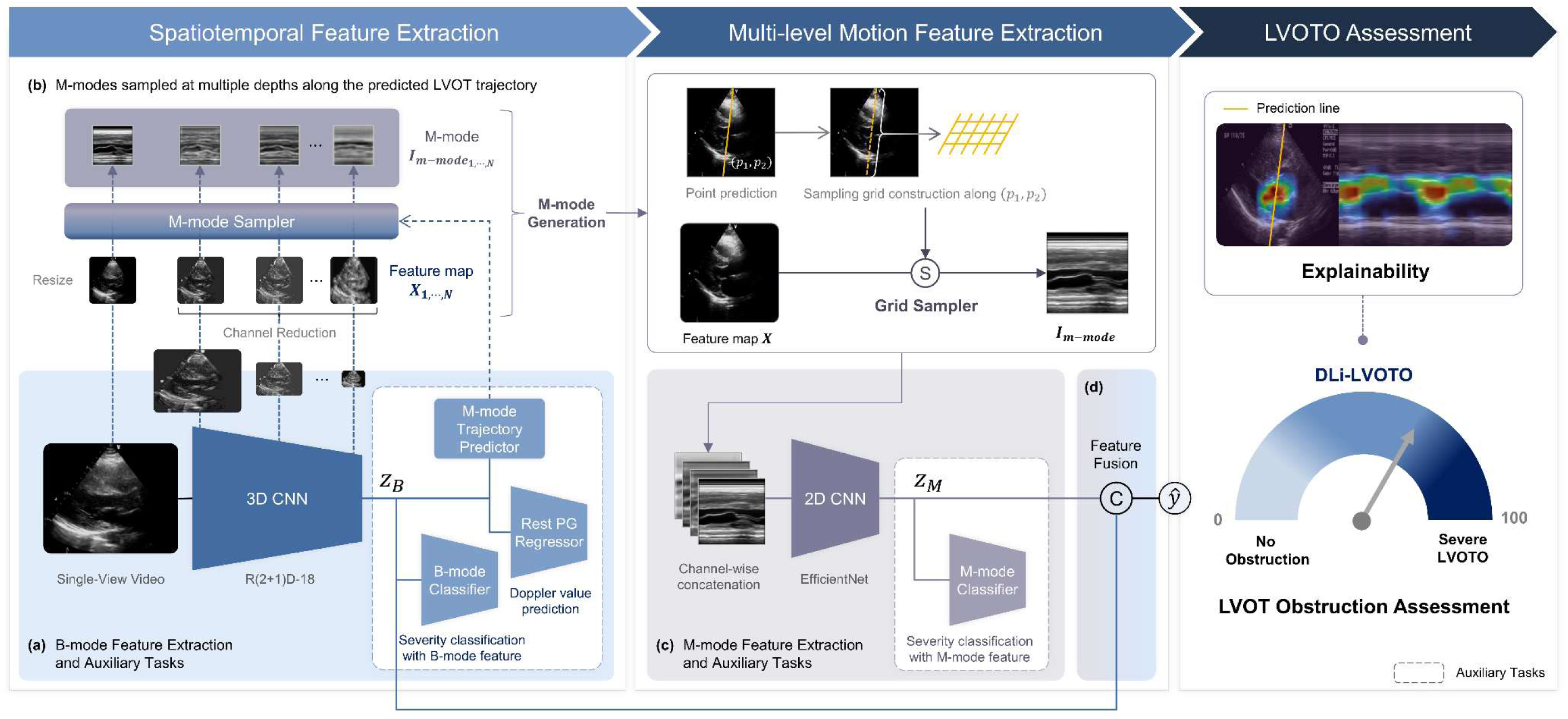
DL Framework for LVOT Obstruction Assessment. CNN, convolutional neural network; DL, deep learning; LVOT, left ventricular outflow tract; LVOTO, left ventricular outflow tract obstruction; PG, pressure gradient.

Our network architecture employs a modified R(2+1)D-18 backbone,^15^ based on a ResNet-18^16^ using factorized 3-dimensional convolutions (2D spatial + 1D temporal) to retain full temporal resolution for video data. To effectively capture the motion dynamics critical for predicting severe LVOTO, we introduced an automated M-mode generation process that leverages a spatial transformer network (STN).^17^ The model autonomously learns and predicts an optimal M-mode line and extracts the motion information along that line in a fully differentiable manner. An auxiliary mean squared error (MSE) loss constrains the M-mode trajectory to pass through the mitral valve (MV) tip, ensuring alignment with clinically relevant anatomical landmarks. This approach mimics the conventional clinician-derived M-mode acquisition from the PLAX view, where the M-mode line is typically placed to evaluate LVOT dynamics effectively. Additionally, we implemented multi-level M-mode generation at various network depths, fusing these representations to create comprehensive motion embeddings. The generated M-mode representations were independently processed using a pre-trained EfficientNet-B3^10,18^ M-mode, and spatiotemporal (B-mode) features were then fused via concatenation.

The model was trained using a supervised learning approach with a primary classification task to distinguish severe and non-severe LVOTO, optimized using a binary cross-entropy (CE) loss. Since severe LVOTO is defined based on peak PG, which can be augmented by provocation, but PLAX videos used in this study were recorded at rest, we introduced an auxiliary regression task to predict the LVOT PG measured at rest. This guided the network to learn subtle hemodynamic features from resting images that correlate with LVOTO severity, allowing it to infer additional predictive cues beyond rest-only inputs for a diagnosis that often requires provocation. Additionally, an early exit strategy with auxiliary classifiers attached to intermediate features improved gradient flow and training stability. Overall, the training objective was formulated as a summation of three loss components: binary CE loss for classification, mean absolute error (MAE) loss for PG regression, and anatomical MSE loss to ensure physiologically meaningful feature extraction. All loss terms were equally weighted (1.0). To enhance generalization and robustness of the model, we applied random augmentations including noise injection, sector masking, haze, depth-dependent attenuation, dynamic gain variation, brightness/contrast adjustment, temporal noise, sharpening, and geometric transforms such as translation, rotation, and cropping.^19^

For patient-level DL index of LVOTO (DLi-LVOTO; range 0-100), multiple PLAX videos from a single patient were individually analyzed, and their DLi-LVOTO scores were averaged. Additional details on the model architecture, M-mode processing, and training parameters are available in **Supplemental Methods 1**. Further analyses on the incremental contributions of key components, including auxiliary regression loss, anatomical alignment loss, early-exit strategy, and multi-level M-mode generation, are presented in **Supplemental Methods 2**. The model represents the latest advancement in our artificial intelligence (AI)-driven HCM evaluation module (USfeat_HCM.ai, Ontact Health, Korea), which integrates validated features such as view classification and automatic measurement capabilities.^10,12,13,20^

### 2.4. Model Validation and Statistical Analysis

The performance of our DL-based LVOTO prediction model was validated using an internal test dataset (ITDS) and an independent external dataset (DHDS). Additionally, we assessed the model’s performance in serial TTE examinations of patients who underwent LVOT gradient reduction therapy, including surgical myectomy and Mavacamten treatment.

The DL-based prediction algorithm’s performance for severe LVOTO in the ITDS and DHDS was evaluated using receiver operating characteristics (ROC) curve analysis, with the area under the curve (AUC) and 95% confidence interval (CI) as key performance measures. We also visualized the distribution of the DLi-LVOTO across categories of no or insignificant LVOTO, significant LVOTO, and severe LVOTO using violin plots. Additionally, we stratified DLi-LVOTO into 10-unit intervals and evaluated its diagnostic performance at each cutoff, assessing key performance metrics, including accuracy, F1-score, sensitivity, specificity, positive predictive value (PPV), and negative predictive value (NPV).

To further assess model robustness, subgroup analyses were performed based on HCM phenotypes and PLAX image quality (IQ). HCM was categorized into three phenotypes – septal, diffuse/mixed, and apical subtypes – based on morphological characteristics observed during TTE. In addition, PLAX IQ was classified into three level: Excellent (clear visualization of key structures with sharp endocardial borders), Good (minor shadowing or dropout with adequate diagnostic quality), and Fair (partial obscuration due to artifacts or suboptimal window, yet still interpretable).^21^

Saliency maps were generated using the Gradient-weighted Class Activation Mapping (Grad-CAM)^22^, with representative maps presented for each severity level to highlight the areas with the greatest influence on the model’s prediction. In patients who were treated with surgical myectomy or Mavacamten, we compared baseline and follow-up (FU) TTE studies performed before and after treatment by visualizing changes in LVOT peak PG and DLi-LVOTO.

## 3. RESULTS

### 3.1. Baseline Clinical and Echocardiographic Characteristics

Baseline clinical and echocardiographic characteristics across datasets are shown in **Table 1**. Overall, the DDS exhibited a relatively balanced distribution of apical, septal, and diffuse or mixed types of HCM. However, in the DHDS, the septal type was the most prevalent. Regarding significant LVOTO prevalence, DDS showed a 27-29% prevalence, whereas DHDS had a lower prevalence of 15.4%. Additionally, when comparing the Mavacamten treatment dataset and the surgical myectomy dataset, patients who received Mavacamten were generally older and had a higher diffuse or mixed-type prevalence. In contrast, those who underwent surgical myectomy were relatively younger, with a predominance of the septal type.

**Table 1.** Baseline Characteristics.

|  | Developmental Dataset |  |  | External Test Dataset |  |  |
| --- | --- | --- | --- | --- | --- | --- |
|  | Training Dataset | Validation Dataset | Internal Test Dataset (ITDS) | Distinct Hospital Dataset (DHDS) | Surgical Myectomy Dataset | Mavacamten Treatment Dataset |
| *Demographics* |  |  |  |  |  |  |
| Number of patients | 578 | 71 | 73 | 573 | 17 | 13 |
|  | 65 (55–74) | 60 (51–72) | 65 (56–75) | 60 (50–70) | 60 (46–65) | 69 (51–77) |
| Male | 385 (66.6) | 55 (77.5) | 39 (53.4) | 384 (67.0) | 9 (52.9) | 6 (46.2) |
| HCM type |  |  |  |  |  |  |
| Apical | 226 (39.1) | 24 (33.8) | 27 (37.0) | 114 (19.9) | 0 (0.0) | 0 (0.0) |
| Septal | 178 (30.8) | 24 (33.8) | 20 (27.4) | 409 (71.4) | 10 (58.8) | 3 (23.1) |
| Diffuse or mixed | 161 (27.9) | 23 (32.4) | 25 (34.2) | 40 (7.0) | 7 (41.2) | 9 (69.2) |
| Others | 13 (2.2) | 0 (0.0) | 1 (1.4) | 10 (1.7) | 0 (0.0) | 1 (7.7) |
| *Echocardiographic data* |  |  |  |  |  |  |
| Number of TTEs | 809 | 111 | 87 | 1,334 | 112 | 44 |
| LVEDD, mm | 45 (41–48) | 45 (42–48) | 46 (42–50) | 47 (44–51) | 44 (40–49) | 42 (37–46) |
| IVS, mm | 15 (12–18) | 16 (14–18) | 14 (12–18) | 18 (16–20) | 17 (15–20) | 18 (14–22) |
| LVEF, % | 65 (61–69) | 65 (61–69) | 65 (62–69) | 68 (62–73) | 65 (58–69) | 67 (62–69) |
| LAVI, ml/m<sup>2</sup> | 45 (36–58) | 48 (39–59) | 47 (38–60) | 44 (35–56) | 59 (42–77) | 54 (46–79) |
| E/e | 13 (10–18) | 13 (10–17) | 13 (9–18) | 15 (11–19) | 19 (15–24) | 19 (13–29) |
| LVOTO | 235 (29.0) | 31 (27.9) | 24 (27.6) | 205 (15.4) | 55 (49.1) | 26 (59.1) |
| Significant (30≤ to <50mmHg) | 73 (9.0) | 13 (11.7) | 12 (13.8) | 48 (3.6) | 13 (11.6) | 9 (20.5) |
| Severe (≥50mmHg) | 162 (20.0) | 18 (16.2) | 12 (13.8) | 157 (11.8) | 42 (37.5) | 17 (38.6) |
| SAM | 97 (12.0) | 6 (5.4) | 7 (8.0) | 163 (12.2) | 43 (38.4) | 21 (47.7) |

Values are given as numbers (percentage) or median (interquartile range).
Abbreviations: E/e, ratio of early diastolic mitral inflow velocity to early diastolic mitral annular velocity; HCM, hypertrophic cardiomyopathy; IVS, interventricular septum; LAVI, left atrial volume index; LVEDD, left ventricular end-diastolic dimension; LVEF, left ventricular ejection fraction; LVOTO, left ventricular outflow tract obstruction; SAM, systolic anterior motion; TTE, transthoracic echocardiography.

### 3.2. Performance of DL-based prediction of LVOT obstruction

Our DL-based LVOTO prediction model was able to reliably detect the presence of severe LVOTO using single-view, 2D PLAX videos, achieving an AUROC of 0.97 (95% CI: 0.92-1.00) in ITDS and 0.93 (95% CI: 0.92-0.95) in DHDS. (**Figure 3**) Subgroup analysis based on HCM phenotypes, including septal, diffuse/mixed, and apical subtypes, demonstrated that the model’s performance remained consistently robust across all subtypes in both ITDS and DHDS validation. (**Supplemental Result 1**) Furthermore, IQ subgroup analysis revealed that model performance remained stable regardless of PLAX view quality. Even in Good and Fair IQ subgroups, the AUROC values were comparable to those in the Excellent group, indicating the model’s robustness against variations in IQ (**Supplemental Result 2**) The DLi-LVOTO distribution showed a gradual increase with LVOTO severity in both ITDS and DHDS, reflecting a consistent relationship between the predicted scores and clinical classification (**Figure 3**).

**Figure 3.**
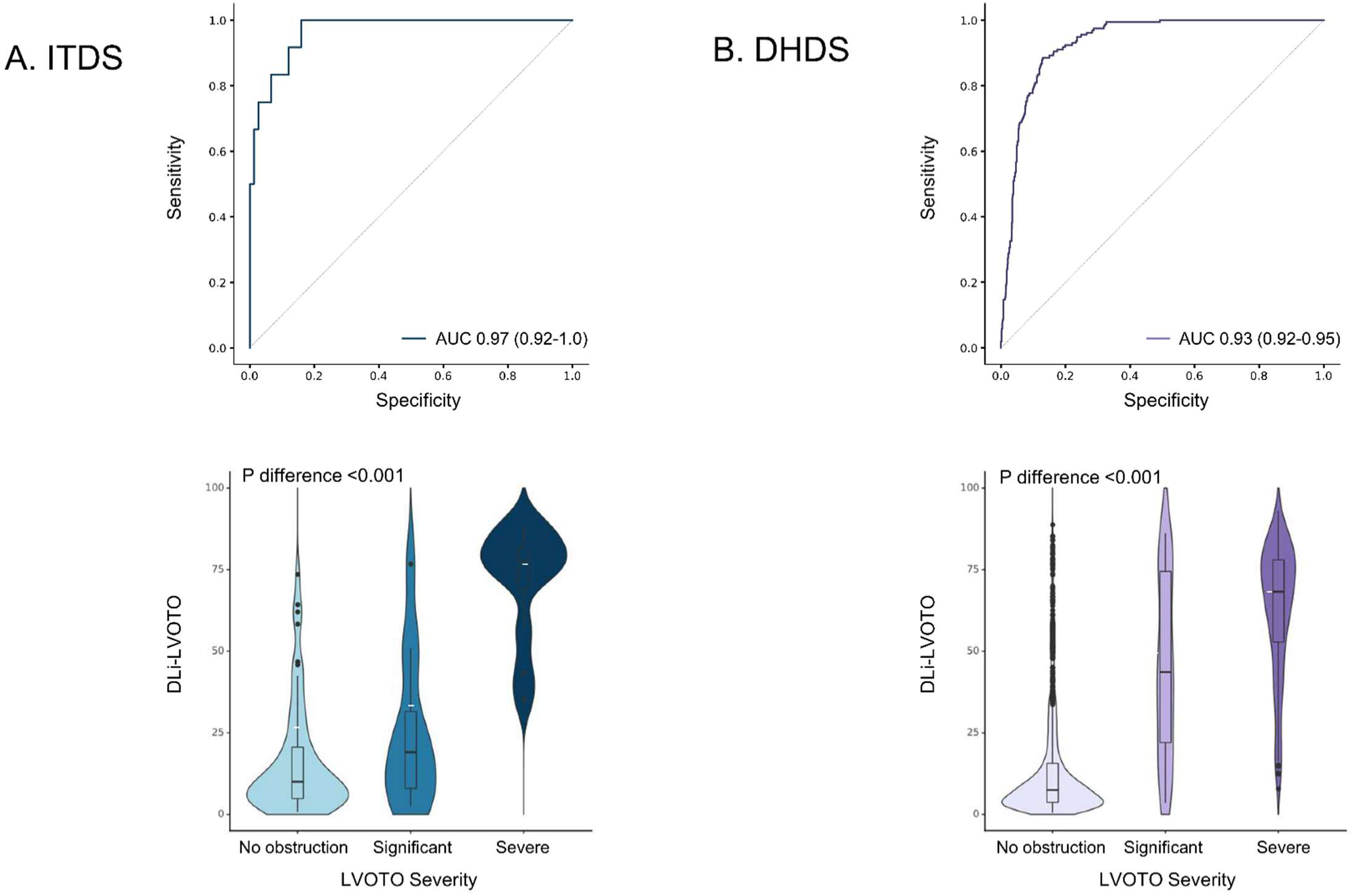
Model performance in the internal and external validation sets. DHDS, distinct hospital dataset; DLi-LVOTO, deep learning index for LVOTO; ITDS, internal test dataset; LVOTO, left ventricular outflow tract obstruction.

**Table 2** presents the diagnostic performance of DLi-LVOTO across different cutoff values for identifying severe LVOTO in both the ITDS and DHDS. In the ITDS, the highest accuracy (94.3%) was observed at a DLi-LVOTO cutoff of 70, demonstrating excellent specificity (97.3%) and NPV (96.1%). This trend was consistently reproduced in the DHDS, where applying the cutoff of 60 yielded an accuracy of 91.3%, with a specificity of 94.6% and an NPV of 95.5%. These results suggest that low DLi-LVOTO scores are strongly associated with the absence of severe LVOTO, effectively allowing the model to serve as a clinical gatekeeper by safely ruling out obstruction. Notably, there is a trade-off between specificity and sensitivity across different cutoff values. While higher cutoffs improve specificity, they are accompanied by reduced sensitivity, potentially leading to missed severe LVOTO cases. Therefore, in the present study, setting a single fixed cutoff may be challenging.

**Table 2.** Diagnostic Performance of DLi-LVOTO Cutoffs for Identifying Severe LVOTO.

| DLI-LVOTO<br>Cutoff | Internal Test Dataset (ITDS) |  |  |  |  |  | Distinct Hospital Dataset (DHDS) |  |  |  |  |  |
| --- | --- | --- | --- | --- | --- | --- | --- | --- | --- | --- | --- | --- |
|  | Accuracy<br>(%) | F1-score | Sensitivity<br>(%) | Specificity<br>(%) | PPV<br>(%) | NPV<br>(%) | Accuracy<br>(%) | F1-score | Sensitivity<br>(%) | Specificity<br>(%) | PPV<br>(%) | NPV<br>(%) |
| **10** | 55.2 | 38.1 | **100.0** | 48.0 | 23.5 | **100.0** | 65.4 | 40.3 | **99.4** | 60.8 | 25.3 | **99.9** |
| **20** | 73.6 | 51.1 | **100.0** | 69.3 | 34.3 | **100.0** | 78.7 | 50.7 | 93.0 | 76.8 | 34.8 | 98.8 |
| **30** | 83.9 | 63.2 | **100.0** | 81.3 | 46.2 | **100.0** | 84.4 | 57.7 | 90.4 | 83.6 | 42.4 | 98.5 |
| **40** | 87.4 | 66.7 | 91.7 | 86.7 | 52.4 | 98.5 | 87.6 | 61.5 | 84.1 | 88.1 | 48.5 | 97.6 |
| **50** | 90.8 | 71.4 | 83.3 | 92.0 | 62.5 | 97.2 | 89.5 | 63.5 | 77.7 | 91.1 | 53.7 | 96.8 |
| **60** | 92.0 | 72.0 | 75.0 | 94.7 | 69.2 | 95.9 | **91.3** | **64.2** | 66.2 | 94.6 | 62.3 | 95.5 |
| **70** | **94.3** | **78.3** | 75.0 | 97.3 | 81.8 | 96.1 | 90.5 | 53.1 | 45.9 | 96.4 | **63.2** | 93.0 |
| **80** | 92.0 | 58.8 | 41.7 | **100.0** | **100.0** | 91.5 | 89.1 | 29.8 | 19.7 | **98.3** | 60.8 | 90.2 |

Abbreviations: NPV, negative predictive value; PPV, positive predictive value.

For each severity level, we present representative samples with Grad-CAM saliency maps overlaid on PLAX views, specifically highlighting the LVOT region (**Figure 4** and **Supplemental Video 1**). These results demonstrate that our model accurately identifies the relevant regions for evaluating LVOTO across all severity levels without supervision.

**Figure 4.**
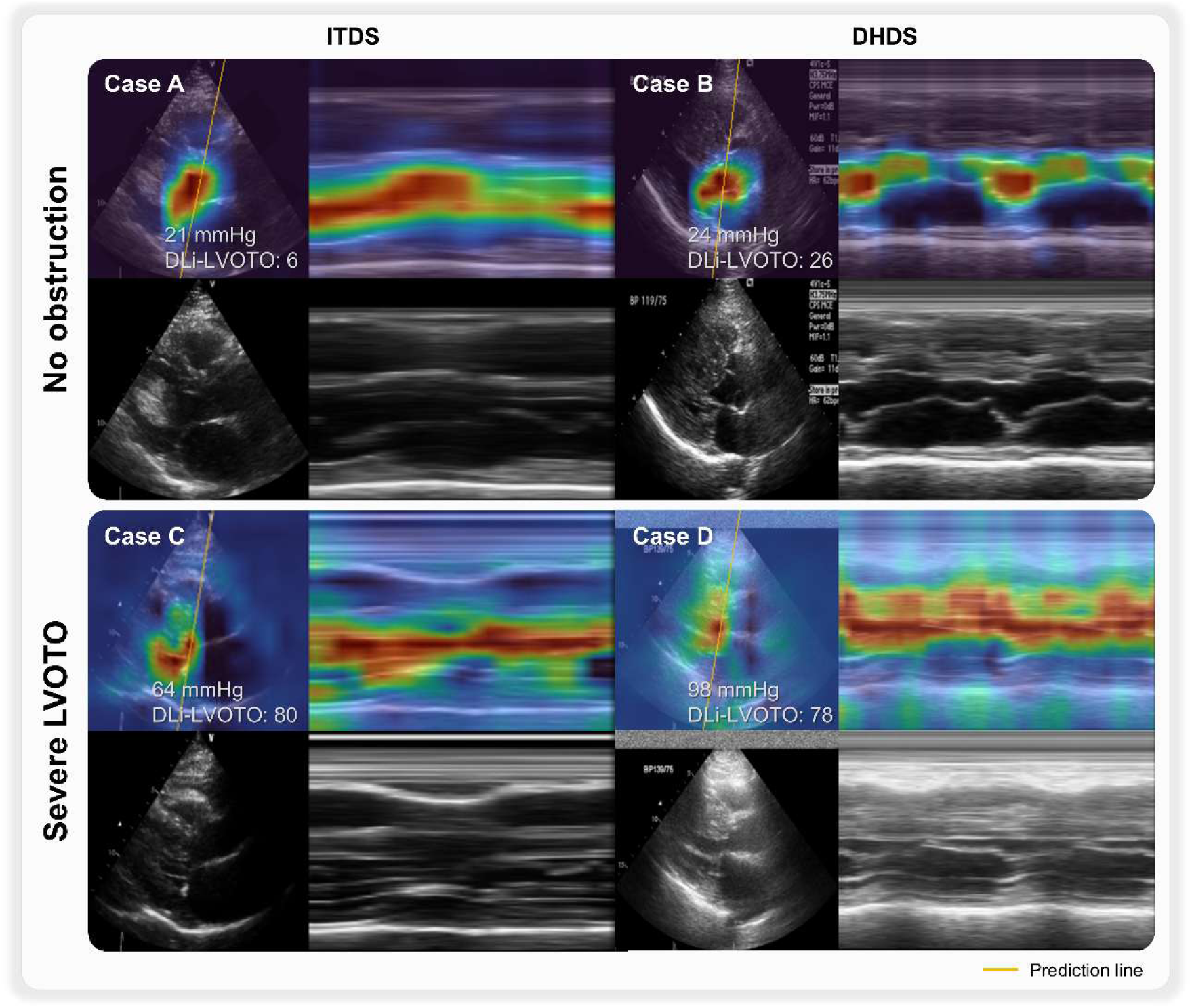
Explainability Analysis for DLi-LVOTO using Saliency Map. DHDS, distinct hospital dataset; DLi-LVOTO, deep learning index for LVOTO; ITDS, internal test dataset; LVOTO, left ventricular outflow tract obstruction.

### 3.3. DLi-LVOTO Changes After LVOT Gradient Reduction Treatment

In patients who underwent LVOT gradient reduction treatment, we visualized the changes in LVOT peak PG and DLi-LVOTO between baseline and FU TTE exams (**Figure 5**). In patients who underwent surgical myectomy, LVOT peak PG showed a marked reduction post-surgery, accompanied by a corresponding decrease in DLi-LVOTO. Similarly, in patients treated with Mavacamten, both LVOT peak PG and DLi-LVOTO progressively decreased over the course of treatment compared to baseline. Representative cases are presented in **Figure 6**. In the patient who underwent surgical myectomy, a significant reduction in peak PG was accompanied by a corresponding decrease in DLi-LVOTO. In a while, for the patient treated with Mavacamten, when the initial treatment response was insufficient, both peak PG and DLi-LVOTO remained elevated. However, with dose escalation, a gradual decrease in both parameters was observed. To further investigate the relationship between treatment-induced changes in peak PG and DLi-LVOTO, we visualized these changes at the individual case level (**Supplemental Result 3** and **4**). The results demonstrated that changes in DLi-LVOTO closely paralleled changes in LVOT peak PG following treatment.

**Figure 5.**
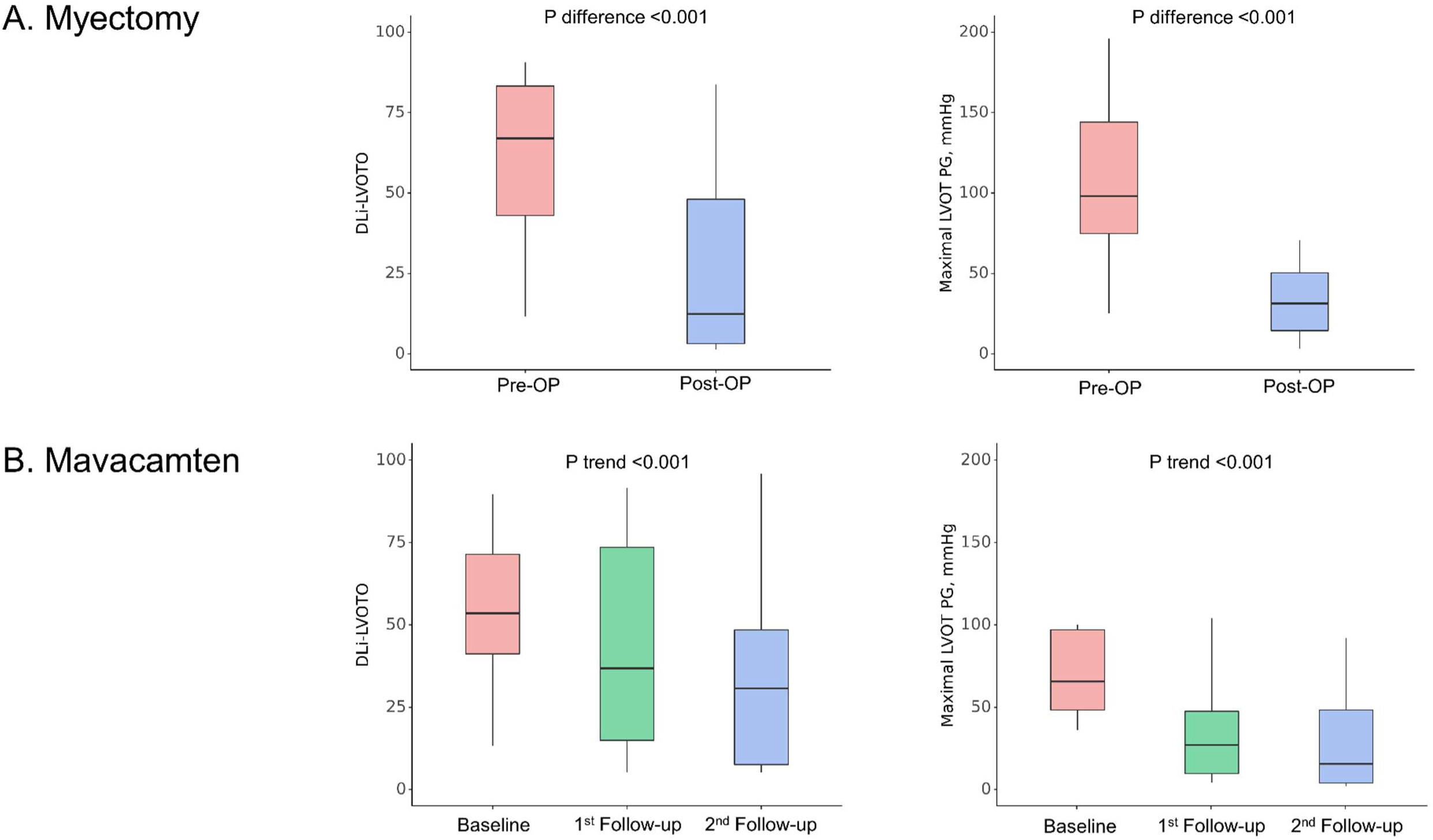
Interval changes in DLi-LVOTO and peak PG after LVOT Gradient Reduction Treatment. DLi-LVOTO, deep learning index for LVOTO; LVOT, left ventricular outflow tract; PG, pressure gradient.

**Figure 6.**
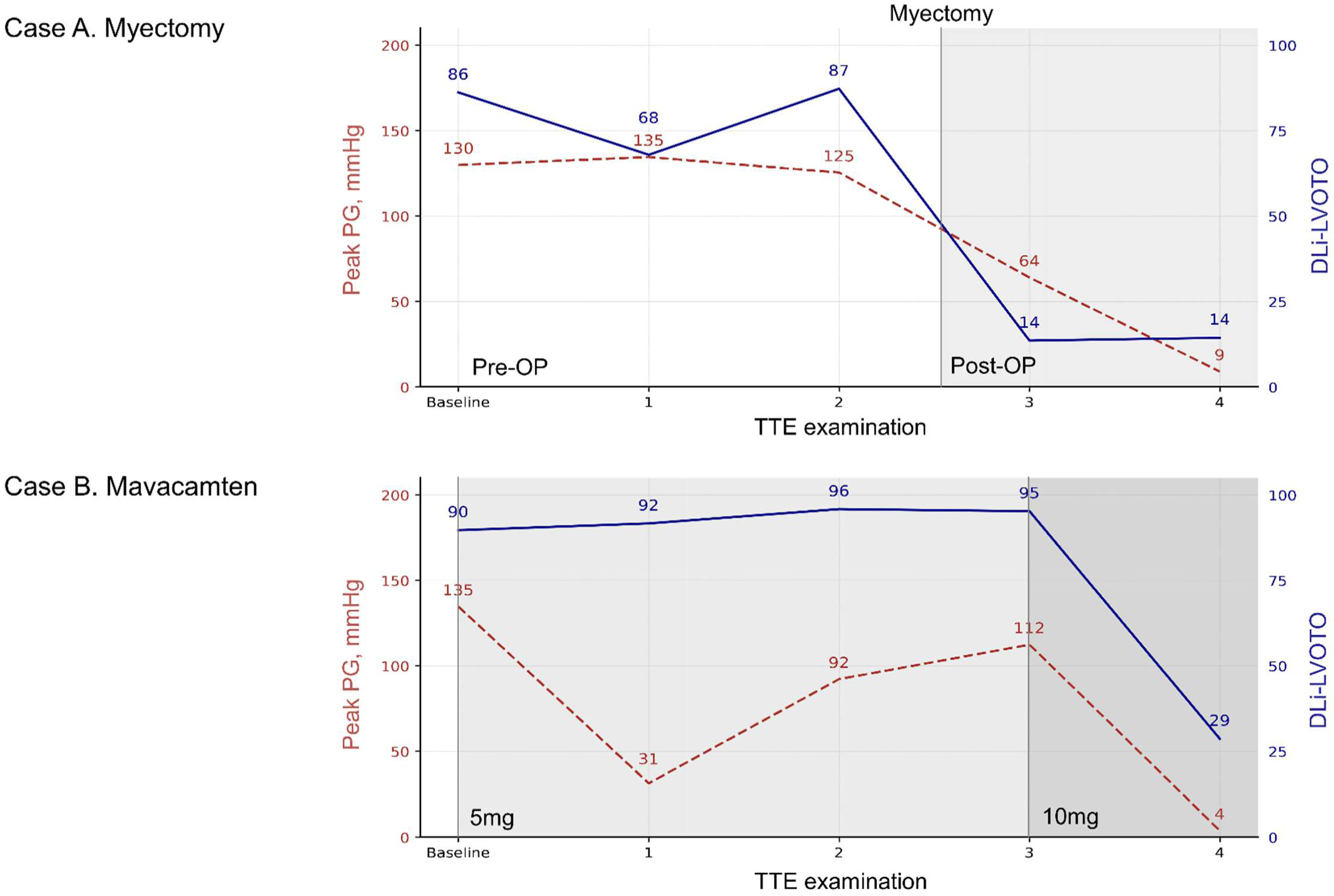
Representative Cases for Trends in DLi-LVOTO and PG with LVOT Gradient Reduction Treatment. DLi-LVOTO, deep learning index for LVOTO; LVOT, left ventricular outflow tract; PG, pressure gradient.

## 4. DISCUSSION

We have developed and validated a DL-based model for predicting severe LVOTO in HCM using only a single TTE view. Despite relying solely on the PLAX view, the model demonstrated high accuracy in predicting severe LVOTO, with robust performance validated not only in the ITDS but also in independent external datasets. These findings suggest that this approach could be a reliable and efficient alternative to conventional methods. Furthermore, we observed that changes in LVOT peak PG following treatment correlated with corresponding changes in DLi-LVOTO within the same patient, highlighting the model’s potential utility in tracking treatment response.

The application of AI in echocardiography has advanced significantly over the past decade. Early research focused on automating manual measurements, including view classification, target structure segmentation, and quantification. However, one of the core aspects of echocardiographic examination – the expert-driven visual analysis – remained largely unchallenged by AI. Recent DL models have begun to mimic expert eyeball analysis, predicting diastolic dysfunction from an A4C view and AS using PLAX or parasternal short-axis views.^4–7,23^ These models do not aim for a standalone definitive diagnosis but rather serve as decision-support tools, flagging potential disease and guiding further workup. Our DLi-LVOTO model extends this paradigm by enabling real-time LVOTO risk estimation during routine B-mode scanning. It could be integrated into handheld ultrasound devices that lack Doppler capability, allowing early detection and timely evaluation in resource-limited settings. Additionally, it can be incorporated into the standard echocardiography workflow, generating the DL-derived index during initial PLAX acquisition. This real-time feedback would prompt targeted Doppler evaluation with provocation maneuvers for high-risk cases, streamlining the diagnostic process. DLi-LVOTO also promises longitudinal monitoring during treatment, tracking changes to evaluate the effectiveness of Mavacamten therapy or septal myectomy. To fully validate these applications, larger-scale prospective studies focusing on both the diagnostic effectiveness of DLi-LVOTO integration and treatment response tracking are warranted.

Although predicting LVOTO and AS from B-mode TTE video may be similar, key differences exist. Unlike AS, which is typically a fixed obstruction, LVOTO is inherently dynamic, fluctuating throughout the day and influenced by physiological and hemodynamic conditions. Therefore, accurate LVOTO assessment with TTE often requires provocation maneuvers, such as the Valsalva maneuver, or stress tests to induce obstruction and unmask its severity. This variability makes predicting LVOTO using only resting B-mode videos challenging. However, certain structural and hemodynamic factors that predispose patients to LVOTO development can still be evaluated on resting TTE. These include a narrowed LVOT, sigmoid septum morphology, systolic anterior motion (SAM) of the MV, and small LV cavity size, all of which contribute to flow acceleration and obstruction under specific conditions. Based on this understanding, we hypothesized that the PLAX view – a foundational TTE perspective – could serve as an optimal input for DL-based LVOTO prediction. Its clear visualization of the LVOT and key anatomical determinants of obstruction makes it particularly suited for this purpose. As a result, we successfully developed a DL-based model trained exclusively on single-view video data, capable of reliably identifying severe LVOTO, even without Doppler input or multi-view analysis. This distinguishes our approach from the only existing study (preprint, not peer-reviewed) that attempted to predict LVOTO using A4C views alone.^8^ We believe this difference in anatomical visualization is one of the key factors contributing to the enhanced performance observed in our study.

Several previous studies on echocardiographic disease prediction have adopted different strategies, each with inherent limitations. For example, Huang et al.^23^ extracted single frames from echocardiogram videos, converted them to grayscale, and resized them to 64×64 pixels before training a WideResNet-28 for view classification and AS diagnosis. While this 2D method is computationally efficient, its static nature makes capturing the dynamic motion patterns critical for conditions like LVOTO challenging. In contrast, Holste et al.^4^ employed 3D convolutional neural networks (CNNs) with extensive self-supervised pretraining and deep ensemble methods, assuming that temporal patterns would emerge implicitly from the video data. Although such approaches benefited from modeling spatiotemporal information, they did not explicitly focus on the motion of key anatomical structures and were computationally expensive. Similarly, our group previously utilized an R(2+1)D architecture with a continuum-aware multi-task loss to accurately classify and comprehensively assess the AS continuum.^5,6,24^ However, this method still relied on the network to implicitly learn temporal dynamics rather than explicitly track motion. In contrast, our current LVOTO prediction model was explicitly designed to capture both morphological and dynamic motion features crucial for LVOTO assessment. By incorporating multi-level M-mode generation and embeddings, our method enhances the detection of subtle transient motion, such as the SAM of the MV, and their resulting hemodynamic consequences. These architectural advancements contributed to the superior performance compared to a prior attempt to detect LVOTO from B-mode video only^8^, indicating its robustness and potential for clinical translation of our model.

While B-mode-derived M-mode representations have been explored in AI-driven echocardiographic analysis, the prior approach has limitations. For instance, a previous study attempted to improve cardiac function prediction by incorporating M-mode imaging for ejection fraction estimation.^25^ However, their method relied on fixed sampling lines selected via heuristic rules, producing static M-mode representations that could not adapt to individual patient anatomy or dynamic motion patterns. Additionally, because M-mode extraction was performed as a separate preprocessing step, the model could not refine feature selection end-to-end manner, potentially limiting diagnostic performance. Our approach fundamentally differs by incorporating M-mode generation directly within the DL framework through the spatial transformer module. This allows the network to autonomously determine the most informative M-mode trajectory on a frame-by-frame basis, optimizing motion extraction as part of the training process. By jointly optimizing the trajectory selection, feature extraction, and classification, our model overcomes the constraints of the static M-mode placement and enables more robust LVOTO detection. Furthermore, our model enhances feature extractions through multi-resolution processing and bilinear fusion of B-mode and M-mode representations, providing a comprehensive understanding of both structural morphology and motion dynamics within an end-to-end framework. This integration improves classification accuracy and enhances clinical interpretability, as the network autonomously identifies and prioritizes diagnostically relevant features directly from the data. Importantly, our automated M-mode generation yields visualizations that align with traditional clinical practice, where M-mode imaging is highly valued for capturing motion over time by producing M-mode images that display critical dynamic features in a format familiar to clinicians. Moreover, Grad-CAM overlays on these generated M-mode images further enhance interpretability by highlighting the precise regions and moments of abnormal motion that drive the network’s predictions.

Despite the robust performance demonstrated across internal and external datasets, it is important to recognize the trade-off between specificity and sensitivity observed at different cutoff values. As noted in our analysis, higher cutoffs increase specificity but simultaneously reduce sensitivity, potentially leading to missed severe LVOTO cases. This trade-off highlights the need for careful consideration when selecting an optimal threshold, particularly in clinical settings where the balance between false positives and false negatives must be managed according to diagnostic priorities. Unlike fixed Doppler-based criteria, our DL-based model offers the flexibility to adjust cutoff values depending on clinical context — for example, prioritizing sensitivity in initial screenings or specificity in pre-treatment evaluations. Further studies are warranted to explore optimal threshold strategies tailored to specific clinical scenarios.

This study has several limitations. Although we developed and rigorously validated our DL-based model using multi-center data, including internal and external validation, all datasets were retrospectively collected from tertiary centers in South Korea. As a result, caution is required when interpreting the findings and applying them to clinical practice. Further validation across diverse populations and healthcare settings is essential to enhance generalizability. Additionally, while the DL model was evaluated using TTE data from multiple institutions, its performance in resource-limited environments or when used by novice operators remains uncertain. Whether DLi-LVOTO will perform reliably on TTE images acquired in such settings is yet to be determined. However, given that the PLAX view is one of the most fundamental TTE views and is more likely to be adequately obtained than a complete multi-view TTE examination, this suggests that DLi-LVOTO could provide a reliable assessment of LVOTO in HCM patients even in less advanced settings. Secondly, LVOTO is not exclusive to HCM patients and can occur in various clinical settings, such as hyperdynamic states and certain cardiac structural variations. Additional studies are required to determine whether DLi-LVOTO can accurately detect LVOTO in non-HCM patients. Thirdly, this study also examined the changes in DLi-LVOTO in response to Mavacamten or surgical myectomy treatment alongside LVOT PG, although the analysis was conducted on a relatively small patient cohort. Despite the limited sample size, we observed significant difference in DLi-LVOTO before and after treatment, suggesting that our approach is effective in capturing hemodynamic improvements. However, larger scale studies are needed to confirm the reproducibility of these observations. Lastly, future research should explore whether DLi-LVOTO can predict clinical outcomes in HCM patients. Addressing these aspects will be crucial for further validating the clinical utility of this DL-based approach.

In conclusion, our DL-based approach enables the prediction of severe LVOTO using only the PLAX view from TTE, providing a complementary tool in situations where acquiring multiple views or Doppler-based LVOT pressure gradients is challenging. Additionally, DLi-LVOTO may aid in monitoring treatment response, offering insights into hemodynamic changes following surgical myectomy or Mavacamten therapy. This method has the potential to enhance LVOTO evaluation in select clinical scenarios, supporting rather than replacing traditional assessment methods.

## Data Availability

The AI-Hub data may be accessible upon proper request and after approval of a proposal. Data from TDDS cannot be made publicly available due to ethical restrictions set by the IRB of the study institution; i.e., public availability would compromise patient confidentiality and participant privacy. Please contact the corresponding author to request the minimal anonymized dataset. Researchers with additional inquiries about the deep learning model developed in this study are also encouraged to reach out to the corresponding author.

## Contributors

All authors contributed equally to this study. All authors have read and approved the final version of the manuscript. J.K, J.P, J.J, and Y.E.Y verified the underlying data of the current study.

## Acknowledgements

This work was supported by a grant from the Institute of Information & communications Technology Planning & Evaluation (IITP) funded by the Korea government (Ministry of Science and ICT) (No.2022000972, Development of a Flexible Mobile Healthcare Software Platform Using 5G MEC); and the Medical AI Clinic Program through the NIPA funded by the MSIT. (Grant No.: H0904-24-1002).

